# Recruitment of Older African Americans in Alzheimer’s Disease Clinical Trials Using A Community Education Approach

**DOI:** 10.1101/2020.07.16.20155556

**Authors:** Ashley R. Shaw, Jaime Perales-Puchalt, Todd Moore, Patricia Weatherspoon, Melissa Robinson, Carl V. Hill, Eric D. Vidoni

**Affiliations:** Department of Neurology, University of Kansas Medical Center, Kansas City, United States; University of Kansas Alzheimer’s Disease Research Center, University of Kansas Medical Center, Fairway, United States; Black Health Care Coalition, Kansas City, United States; Alzheimer’s Association, Chicago, United States

**Keywords:** Alzheimer’s disease, community-based education, recruitment, African Americans, minority health, clinical trials

## Abstract

African Americans are disproportionately affected by Alzheimer’s disease and related dementias (ADRD) and are two times more likely to develop ADRD compared to their White counterparts. Despite the higher prevalence of ADRD among older African Americans, recent estimates suggest research enrollment by those who identify as African American remains limited. The purpose of the study is to 1) explore how a culturally tailored community education program impacts clinical trial interest and enrollment in ADRD research studies and to 2) identify how applicable the African American community perceived the culturally tailored curriculum. Using a community-engaged research approach, we collaborated with predominately African American serving community-based organizations to support content development and delivery of Aging with Grace (AWG), a culturally tailored ADRD educational curriculum. A total of five AWG presentations were given to 66 attendees. Most attendees (67%) expressed interest in participating in clinical trials after attending AWG. Enrollment increased within an observational study (84%) and lifestyle prevention clinical trials (52%) from 2018 to 2019. Attendees (32%) also perceived an increase in ADRD knowledge from attending AWG and 89.1% believed more African Americans should participate in research. Our work demonstrates the effectiveness of a culturally tailored community education program to enhance knowledge, clinical trial interest, and recruitment into observational studies and lifestyle ADRD clinical trials among older African Americans. Education programs developed in partnership with the community can serve as bridge to research participation for under-represented minorities in clinical research. Future studies should assess long-term retention of knowledge and research readiness.

## Background

It is estimated that 5.8 million Americans had Alzheimer’s disease or a related dementia (ADRD) in 2019 (Association, 2019). African Americans are disproportionately affected by ADRD and are two times more likely to develop ADRD compared to their White counterparts (Association, Thies, & Bleiler, 2013; Gurland et al., 1999; Potter et al., 2009; Rajan, Weuve, Barnes, Wilson, & Evans, 2019). Additionally, minorities including African Americans are often diagnosed at later stages of ADRD, resulting in higher levels of chronic morbidity (i.e. cardiovascular disease, stroke, diabetes) (Zuckerman et al., 2008). Previous research has indicated that variations of lifestyle (e.g. nutrition and exercise) and socioeconomic factors (i.e. access to health care, built environment, education) likely contribute to the disproportionate prevalence and risk of ADRD among African Americans (Prevention, 2020; Yaffe et al., 2013). Specifically, health conditions which are heavily influenced by lifestyle, such as diabetes and cardiovascular disease, are more prevalent among African Americans, and are linked to ADRD risk (Glymour & Manly, 2008; Lines, 2014).

Despite the higher prevalence of ADRD among older African Americans, clinical research participation in this population remains low (Darnell, McGuire, & Danner, 2011; Schnieders, Danner, McGuire, Reynolds, & Abner, 2013). One recent meta-analysis found only 2.4% of ADRD trial enrollees identified as Black or African American (Vyas, Raval, Watt, & Tang-Wai, 2018). Lower participation rates among African Americans have been noted across several conditions, including stroke, and cardiovascular disease (Braunstein, Sherber, Schulman, Ding, & Powe, 2008; Gorelick, Harris, Burnett, & Bonecutter, 1998). Underrepresentation in ADRD clinical research reduces generalizability and ability to accurately assess effectiveness for a population with high disease burden (Levkoff & Sanchez, 2003).

Researchers who have struggled to engage, recruit, and retain racial/ethnic minorities have created labels such as “difficult to access” and “hard to reach”, which inherently shifts blame to the community. However, it is the approach by the clinical research enterprise that reinforces and perpetuates social inequality seen within clinical research today (Vollmer, Osborne, Hertert, & Buist, 1994; Wilkins & Alberti, 2019). Moreover, increasing public awareness and discussion of the exploitation of vulnerable populations for “medical discovery”, including the Tuskegee syphilis (Rockwell, Yobs, & Moore, 1964) and other sexually transmitted disease experiments in Guatemala, (Spector-Bagdady & Lombardo) the cloning of Henrietta Lacks’ cells,(Khan, 2011) radiation exposure and gynecologic experimental surgeries conducted on enslaved African Americans and Native Americans, (Vernon, 2019) has reinforced the current community recognition that medical care and medical research is inequitable (Scharff et al.). Mistrust in research among African Americans is sustained due to inequities in health care systems (e.g. social, economic, environmental, structural racism, and classism) that produce health inequalities (National Academies of Sciences & Medicine, 2017). Similarly, clinical trial designs create multifactorial barriers to participation in research among African Americans (Gilmore-Bykovskyi, Jackson, & Wilkins, 2021; Griffith, Bergner, Fair, & Wilkins, 2021). These barriers include but are not limited to cultural barriers, economic barriers, study design eligibility criteria, lack of awareness, lack of invitation, accessibility to research site location, transportation, and implicit bias in which there may be a perception that racial/ethnic minorities do not want to participate in research (Harris, Gorelick, Samuels, & Bempong, 1996; Hughes, Varma, Pettigrew, & Albert, 2017). It is important to note that willingness to participate in research has been found to be just as high among racial/ethnic minorities as compared to Whites, suggesting that factors other attitudes and beliefs play a greater role in lack of participation in clinical trials (Wendler et al., 2006).

Under-representation of older African Americans in ADRD clinical trials poses a number of threats to external validity, generalizability of research findings and advancements in treatments for minorities, (Hill, 2019) and creates an inability for researchers to develop a comprehensive understanding of why differences in burden of ADRD exist (Singh, Azuine, & Siahpush, 2012). Therefore, it is critical to incorporate strategies that address barriers and structural inequities to achieve more inclusive clinical research. The recent National Plan for ADRD Recruitment (Aging, 2018) and the Alzheimer’s & Dementia Outreach, Recruitment & Engagement Resources website (ADORE) (Aging, 2020) outline strategies and compile materials to support inclusive enrollment into ADRD clinical trials. These strategies emphasize building sustainable community partnerships, considering diversity early in study design, and understanding regional cultural customs.

Researchers have indicated that collaborating with community partners may significantly enhance the effectiveness and retention of minorities in clinical research. For example, in one systematic review, investigators examined the efficacy of recruitment strategies used to enhance enrollment among minorities in clinical trials and concluded that active involvement of community partners was an important factor in the enrollment process (Yancey, Ortega, & Kumanyika, 2006). Previous research has found that culturally tailored, and user-designed (Altman, Huang, & Breland) community education programs, delivered in conjunction with trusted community organizations, can increase research participation among racial/ethnic minorities (Perales-Puchalt et al., 2019; Perales et al., 2018). Community partnerships can be an integral part of clinical research, acting as a gateway to the community, and serving as a tool to address recruitment barriers related to inequitable and untrustworthy research design and implementation. Moreover, community partners provide social endorsement, known physical space, up-to-date understanding of community needs and wishes, and expert understanding of how research may be integrated with other services and supports within the population of interest.

It is important to note that culturally tailored educational curricula have been developed and disseminated in the African American community to enhance knowledge, influence behavior change, and to increase participation in research. A faith-based culturally tailored education program to improve self-care activities related to hypertension among African Americans was shown promising in improving medication adherence (Harvin, Winter, Hoover, & Lewis, 2020). Using a community based participatory research approach, a religiously tailored HIV tool kit (i.e. sermons, print testimonials, response readings) was developed to increase HIV testing rates among African Americans (J. Berkley-Patton et al., 2010; J. Y. Berkley-Patton et al., 2019). Cunningham-Erves et al. (2021) examined the effect of a culturally tailored educational program to enhance participation in cancer trials among African Americans and Latinos. Results demonstrated that the culturally tailored curriculum increased willingness to participate in research and improved trust between the researchers and the community (Cunningham-Erves et al., 2021). This paper expands on the current literature around culturally tailored education by focusing on a disease that disproportionally impacts the African American community. Furthermore, this paper includes additional measures (e.g. open-ended questions) to gain a deeper understanding of the African American community’s perception of the curriculum and needs to inform future community based research.

### Purpose

Given the need for diverse and inclusive clinical trial enrollment and participation, it is imperative we conduct representative research by developing culturally sound recruitment strategies designed to address barriers and facilitators for clinical trials among older African Americans to increase the level of involvement and participation in ADRD clinical research trials. The present study sought to 1) explore how a culturally tailored community-based recruitment method impacts clinical trial interest and enrollment in ADRD research studies and to 2) identify how applicable a Midwestern African American community perceived the culturally tailored curriculum. Our guiding hypothesis was that recruitment strategies developed in partnership with racial/ethnic communities would increase enrollment in ADRD observational, treatment, and prevention research.

## Methods

Using a community-engaged research approach, we collaborated with two Midwestern, predominately African American-serving, organizations (a health advocacy organization and a faith-based organization) to understand community needs and to support the content development of Aging with Grace (AWG); a culturally tailored ADRD educational curriculum. We also collaborated with leaders in these organizations to develop our protocol of questionnaires and activities to capture knowledge, beliefs, and interest in clinical research when delivering AWG presentations. This study was approved by the University of Kansas Institutional Review Board as minimal risk education research (STUDY#2979). All adults over the age of 21 were eligible to participate. Though ADRD are predominant diseases impacting individuals over 60 years old, community members of all ages can serve as sources of community information, and many are eligible to participate in various trials including those for caregiving.

### Development of Aging with Grace

AWG is an ADRD knowledge didactic presentation designed with a primary goal to enhance knowledge of ADRD and ADRD risk reduction. Aligned with our previous work, we developed AWG by first developing the curriculum in partnership with organizations (Perales-Puchalt et al., 2019; Perales et al., 2018). Specifically, we employed user-centered design, (Altman et al.) which has been previously shown to be effective in the development and implementation phases of health-related interventions (Altman et al., 2018). User-centered design is an approach that incorporates the inclusion of collaborative multidisciplinary teams, use of action-oriented models, and the attitudes and opinions of actual users (Ferreira, Song, Gomes, Garcia, & Ferreira, 2015; Roberts, Fisher, Trowbridge, & Bent, 2016).

For AWG we first identified factors related to ADRD knowledge among minorities by conducting a literature review and developing a draft curriculum. Second, we solicited feedback from the research team, leadership of community partners, and ADRD experts including neurologists, dieticians, exercise specialists and researchers as we shaped the curriculum. Lastly, after constructing an initial draft, we presented AWG to community members at two presentation series, incorporating further suggestions, prior to the work presented here.

Considering the contexts in which we intended to use AWG, we further refined the curriculum following the Cultural Accommodation Model (CAM), which applies culture-specific constructs to adequately address emerging needs for diverse communities (Burrow-Sánchez, Minami, & Hops, 2015; Perales et al., 2018). For the present study, CAM provided a foundation to the social context in which the AWG content was developed, organized, structured, and delivered in a culturally appropriate manner. Using CAM principles, the research team incorporated several culture-specific variables such as embedding scriptures for religious and spirituality purposes, (Taylor & Chatters, 2010) culturally specific dietary preferences, educational materials designed in a manner that was simple, colorful, including images and video content of African American community members, (Yang, Buys, Judd, Gower, & Locher, 2013) and use of an interactive approach that emphasized respect and empowerment in the African American community (Utz et al., 2008) (e.g. Insert Figure 1 here).

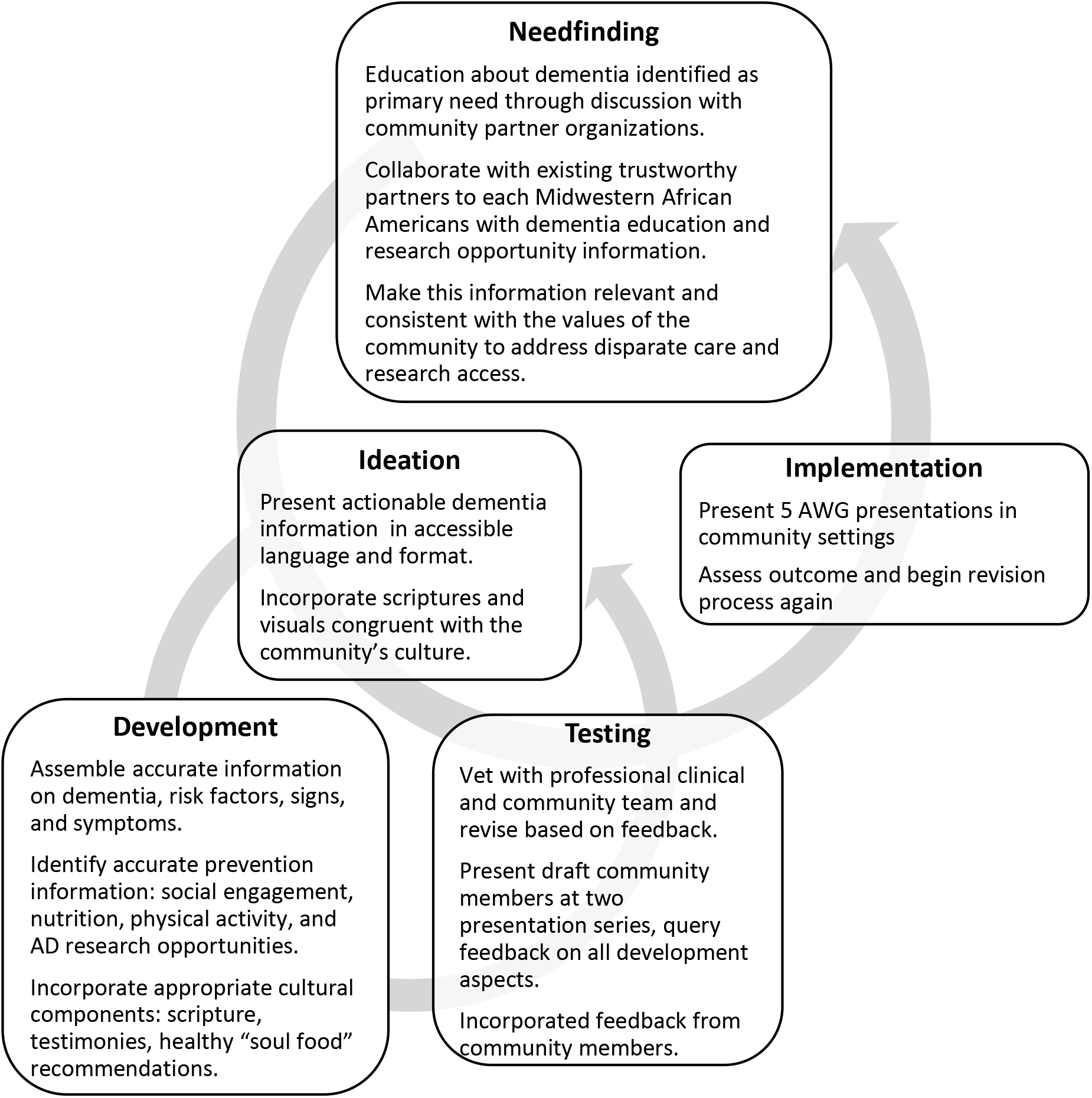

The resulting two-hour presentation included five primary topic areas outlined in Table 1. After developing AWG, we collaborated with predominately African American community organizations, including independent faith communities and our health promotion partner to host a total of five presentations between March and November 2019. Collaboration with these groups assisted in recruitment of attendees and allowed for use of familiar space. (Insert Table 1 here).

**Table 1.** Aging With Grace Topics.

|  |
| --- |
| 1. Dementia risk reduction (social engagement, diet, and exercise) |
| 2. Types of clinical research (e.g. observational, prevention, screening, diagnostic, treatment, and behavioral) |
| 3. History of research within the African American community and current rights and protections as a research participant |
| 4. Importance and need of clinical research in the African American community |
| 5. Currently available research opportunities in the area |

### Measures

#### Demographics

Prior to the start of each AWG presentation, attendees completed an 11-item demographic questionnaire including questions regarding age, gender, education, employment, health status, ADRD family history, work status (e.g. retired, part-time, full-time), cardiovascular comorbidity (e.g. diabetes, high blood pressure, high cholesterol, stroke, heart disease, depression), and race. Race is not biologically anchored, but rather a social construct in which its categorization and measurement in health care research is an evolving discussion (Morning, 2007). We chose to simply ask if individuals identified themselves as “Black” or “African American” and asked a question about their country of origin to better capture cultural identity. At the suggestion of our community partners, we limited the overall number of questions. Attendees completed a questionnaire developed by our team to capture demographics, subjective knowledge of AD, perspectives on research, presentation satisfaction, and research readiness.

#### Subjective Knowledge of AD

We captured knowledge of AD by including questions such as “Do you know anyone with significant memory loss or Alzheimer’s disease?” and “Rate your knowledge of Alzheimer’s disease” (5-item Likert, Very Low [1] to Very High [5]). Following the AWG presentation, attendees again completed the subjective knowledge questionnaire to rate their knowledge of ADRD.

#### Perspectives on Research

We queried interest in study participation upon completion of AWG session (Not interested at all [1] to Very interested [4]). We also asked about participant sentiment towards the statement that “More African Americans should participate in research” (Agreed, felt Neutral, or Disagreed).

#### Presentation Satisfaction

Following the presentation, we measured satisfaction (Poor [1] to Excellent [5]) and applicability to daily life (Not at All [1] to Very applicable [4]) of the AWG curriculum. Using open-ended question format, we asked participants to describe what they liked most about the AWG presentation. Furthermore, in the post survey we asked attendees to identify the top 3 components they liked most about the AWG presentation. Lastly, to assess community needs as it relates to addressing the disproportionate impact of AD on the African American community, we asked participants to identify the top 3 things needed to help fight against AD.

#### Research Readiness

To assess research readiness, we tracked enrollment of attendees in any research study supported at the University of Kansas Alzheimer’s Disease Center (KU ADC) for the period of the one year following the first AWG presentation (Vidoni, Bothwell, Burns, & Dwyer, 2018). As a comparison, we also assessed African American participation in research studies supported by KU ADC in the prior year, March 2018 through February 2019. We did not directly follow AWG participant enrollment into trials as we minimized acquisition of identifiable information at the AWG sessions consistent with partner recommendations.

### Analysis

All data was analyzed using R.(R Development Core Team) Descriptive analyses were performed on demographics, research interest, and research participation. A Wilcoxon signed-rank test was used to compare perceived knowledge before and after the presentation.

## Results

### Participant Demographics

A total of five AWG presentations were held over a 6-month period with an average of 13.2 community members attending each session (n= 66). Most attendees were female (90.5%), between the age of 66-75 (34.9%), retired (69.8%), and had some college education (25.4%). Table 2 shows a summary of the demographic characteristics of AWG participants. (Insert Table 1 here)

**TABLE 2.** Demographics.

| **TABLE 2** |  |
| --- | --- |
| **Demographics** |  |
| **Gender** |  |
| Male | 6 (9.5%) |
| Female | 57 (90.5%) |
| **Age** |  |
| 18-55 | 11 (17.5%) |
| 55-65 | 16 (25.4%) |
| 66-75 | 22 (34.9%) |
| 76 years and older | 14 (22.2%) |
| **Education** |  |
| High School | 9 (12.7%) |
| Vocational/Trade | 4 (6.3%) |
| Some College | 16 (25.4%) |
| Associate | 11 (17.5%) |
| Bachelor | 13 (20.9%) |
| Masters | 10 (15.9%) |
| Doctoral | 1 (1.5%) |
| **Employment Status** |  |
| Employed | 17 (27%) |
| Retired | 44 (69.8%) |
| **Health Conditions** |  |
| Diabetes | 21 (31.8%) |
| Hypertension | 38 (57.6%) |
| High Cholesterol | 26 (39.4%) |
| Stroke | 1 (1.5%) |
| Heart Disease | 6 (9.1%) |
| Depression | 5 (7.6%) |

### Subjective Knowledge and Perception of AWG Curriculum

Most AWG attendees reported that the presentation was high quality (87.9%). Attendees perceived an increase in ADRD knowledge from attending AWG (32%, Z= -4.418, p < 0.001) and 89.1% believed after the educational session that more African Americans should participate in research.

The AWG presentation was rated “Very Good” to “Excellent” with a mean Likert score of 4.45 (range 3-5). The applicability of the presentation was rated ‘Somewhat’ to ‘Very’ Applicable, with a mean Likert score of 3.76 (range 2-4). Attendees indicated that what they liked most about the AWG presentation was that it was informative, interactive, lifestyle focused (diet and exercise modification), and delivered visually (pictures, infographics, and videos).

### Clinical Trials Interest and Enrollment Following AWG

Most attendees (67%) expressed interest in participating in clinical trials after attending an AWG presentation. We therefore assessed inquiries and enrollments of African Americans in KU ADC-sponsored studies before and after the implementation of AWG. The KU ADC maintains a potential participant registry with medical history for future study screening, and tracks enrollments into trials, allowing us to assess enrollment pattern changes (Vidoni et al., 2018). We evaluated enrollment of unique individuals in studies in the one-year period before and after the first AWG presentation in March 2019. We saw changes in clinical trial inquiries and enrollment after offering AWG to Midwestern African Americans. Within our potential participant registry, inquiries increased by 33%, from 63 inquiries in the year prior to AWG to 94 inquires in the year delivering AWG. The KU ADC also enrolled 18 unique African American individuals over a 12-month period out of a total of 273 (6.6%) after AWG, a period that included an extended research shutdown due to the SARS-CoV-2 pandemic. In the year prior to the delivery of AWG, KU ADC enrolled 11 African American individuals out of a total of 462 (2.3%). Of particular note, enrollment of African Americans within our flagship longitudinal observational study increased by 84% following AWG, and enrollment of African Americans in our lifestyle prevention trials increased 52% following AWG.

### Community Needs

Community perceived needs were consistent with recognition of disproportionate impact of ADRD on African Americans. Participants reported education, opportunities for service and care, lifestyle prevention activities as the greatest need for the African American community.

## Discussion

We created a culturally tailored ADRD education curriculum with the community and tested 1) it’s applicability for community members, and 2) clinical study enrollments following delivery. Our results suggest that use of a targeted, community-based recruitment method is effective in enhancing recruitment into ADRD observational and lifestyle trials among older African Americans. This method also improved perceived ADRD knowledge, clinical research participation interest. Despite the common perception that it is difficult to recruit members of racial/ethnic minority (e.g. hard-to-reach) populations into clinical trials, this study demonstrated that dissemination of a culturally tailored ADRD prevention curriculum, can adequately reach and recruit African Americans into clinical trials.

Consistent with the recommendations of Ellad-Gray et al. (2015), one of the key components of this successful recruitment method was partnering with African American community serving organizations, whom were valued and trusted by the African American community (Ellard-Gray, Jeffrey, Choubak, & Crann, 2015). Community partners are essential in terms of serving as gatekeepers to enhance recruitment and can help lessen some of the mistrust towards research, which was demonstrated in this study.

Fisher et al. (2007), encourages use of cultural leverage which is a “focused strategy for improving the health of racial and ethnic communities by using cultural practices, philosophies, or environments” (Johnson, ElbertJAvila, & Tulsky, 2005). In this study cultural leverage was incorporated within both the design and dissemination of the curriculum. Spirituality is an important part of African American culture, therefore the curriculum included scriptures as a means to increase resonance and applicability. Specifically, spiritually serves as a source of meaning and purpose in addition to serving as a framework for understanding and overcoming disparities that impact the African American community. Within this study, ADRD prevention through lifestyle behaviors reflects a spiritual framework that emphasizes “thriving health” for African Americans.

In addition, this study incorporated cultural leverage by disseminating AWG in local community settings (e.g. church, community center), which was shown to be an effective strategy in reaching older African Americans and indirectly served as a bridge to build a relationship between the African American community and academia. It is important to note that cultural leverage addresses racism. Specifically, it addresses institutional racism within academia which has led to reduced opportunities for African Americans to participate in research. Anecdotally, participants particularly appreciated the fact that they could receive ADRD education outside of an academic institution which many perceived as threatening (e.g. due to historical and unethical research practices among underrepresented minorities).

These results are consistent with other studies in which effective recruitment methods for African Americans encompassed a field-based approach involving community outreach and developing partnerships with leaders associated with the African American community in an effort to establish trust with the community (Otado et al., 2015). Also congruent with findings from other studies, we found that engaging the community through education and discussing the nature of the research studies in person was effective in increasing participation (Otado et al., 2015).

### Implications for Practice

Researchers have demonstrated the importance of culturally tailored education design to improve health outcomes and enhance diversity in clinical trials, which has significantly contributed to the field of health disparities (J. Berkley-Patton et al., 2010; Cunningham-Erves et al., 2021; Harvin et al., 2020). However, given the scarcity of culturally tailored community-based recruitment methodology for ADRD among older African Americans, this study provides evidence to support the potential utility of future culturally tailored community-based recruitment methods for ADRD clinical trials. It is important to note that this study did not assess retention in studies, therefore the long-term efficacy of this recruitment method is unknown. Future studies should assess long-term retention of knowledge and research readiness of this AWG community-based recruitment method. Also, participants recruited through AWG enrolled in observational and lifestyle prevention clinical trials, therefore the efficacy of this recruitment method is not understood in terms of recruitment and enrollment into ADRD treatment trials (e.g. investigational medication trials). Additionally, individuals recruited using the AWG community-based recruitment method were from the Midwest, therefore results may not be generalizable to African Americans in other geographical locations.

## Conclusion

In summary, the community-based recruitment method leveraging community partnerships and providing ADRD prevention education can be effective in recruiting older African Americans in ADRD clinical trials. Moreover, authentic communication and collaborative community partnership in research was demonstrated as imperative to the process of community engagement and establishing trust within the African American community. It also suggests that this recruitment method centering on cultural values may have high acceptability among older African Americans. Future research with a larger representative sample size of older African Americans in the United States and assessment of long-term retention is needed to further validate these promising findings.

## Data Availability

All data is available in the attached documents of the manuscript.

## Acknowledgements

Leo and Anne Albert Charitable Trust

Marian Van Dyke and Faith Vidoni Education Fund

